# Reinfection with SARS-CoV-2 and Failure of Humoral Immunity: a case report

**DOI:** 10.1101/2020.09.22.20192443

**Authors:** Jason D. Goldman, Kai Wang, Katharina Röltgen, Sandra C. A. Nielsen, Jared C. Roach, Samia N. Naccache, Fan Yang, Oliver F. Wirz, Kathryn E. Yost, Ji-Yeun Lee, Kelly Chun, Terri Wrin, Christos J. Petropoulos, Inyoul Lee, Shannon Fallen, Paula M. Manner, Julie A. Wallick, Heather A. Algren, Kim M. Murray, Yapeng Su, Jennifer Hadlock, Joshua Jeharajah, William R. Berrington, George P. Pappas, Sonam T. Nyatsatsang, Alexander L. Greninger, Ansuman T. Satpathy, John S. Pauk, Scott D. Boyd, James R. Heath

## Abstract

Recovery from COVID-19 is associated with production of anti-SARS-CoV-2 antibodies, but it is uncertain whether these confer immunity. We describe viral RNA shedding duration in hospitalized patients and identify patients with recurrent shedding. We sequenced viruses from two distinct episodes of symptomatic COVID-19 separated by 140 days in a single patient, to conclusively describe reinfection with a new strain harboring the spike variant D614G. With antibody and B cell analytics, we show correlates of adaptive immunity, including a differential response to D614G. Finally, we discuss implications for vaccine programs and begin to define benchmarks for protection against reinfection from SARS-CoV-2.

## Introduction

The risk of reinfection with SARS-CoV-2 after primary infection has not been consistently demonstrated.^1^ Multiple reports document prolonged viral RNA shedding,^2^ though virus is not likely to be transmissible after 10 days,^3, 4^ or possibly up to 20 days in immunocompromised patients.^5^ These data suggest prolonged shedding of viral remnants, as opposed to ongoing shedding of replication-competent virus. A large case series from the Korean CDC^6^ found lack of transmission events from symptomatic patients repeatedly positive for SARS-CoV-2 after negative testing. Most case reports do not distinguish between prolonged shedding and reinfection.^7-9^ Without viral sequencing analysis, we cannot exclude the possibility that prolonged shedding in some patients may actually be reinfection. Notably, reports from Hong Kong and Nevada describe reinfection 5 and 2 months after primary infection, respectively.^10, 11^

After SARS-CoV-2 infection, most persons develop anti-SARS-CoV-2 antibody responses characterized by rising IgG, IgM and IgA to viral spike, receptor binding domain (RBD) or nucleocapsid (N) antigens.^12^ By 4 weeks after symptoms onset, IgM and IgA decline substantially, as does IgG in patients with mild or asymptomatic infections, while IgG persists at higher levels after severe COVID-19 illness.^13^ Evidence suggests that SARS-CoV-2-specific antibodies can be protective, as indicated by the lack of infection in those with pre-existing neutralizing antibodies (nAbs) in a recent high attack rate outbreak aboard a fishing vessel.^14^ Convalescent plasma programs are based on the assumption that humoral immunity will aid in the response to SARS-CoV-2, ^15^ as are vaccine programs aiming to provide durable herd immunity.^16^ However, correlates of immunity from reinfection have not been established due to the few reinfection. ^10, 11^ Here, we use whole viral genome sequencing to define a new reinfection case. We then present antibody and B cell analyses to evaluate the patient’s lack of immunity against a new SARS-CoV-2 strain.

## Results

### Population sampling

Between March 1^st^ and August 12^th^, 2020, 11,622 patients were tested for SARS-CoV-2 by rt-PCR (**Figure S1**). Of these, 643 patients had at least one positive test (5.5% positivity) and 176 patients had at least two positive samples. Time from first positive to last positive was determined as the shedding duration (**Figure 1A**). The median (interquartile range) shedding duration was 12.1 (6.4, 24.7) days, with a positively skewed distribution (kurtosis = 10.7). Shedding was <59 days in 95% of patients, and was >75 days in only two patients. Re-positivity was observed in 43 patients (**Figure 1B**) with patterns suggesting: 1) inadequate sampling technique, 2) assay limitations with the Ct result hovering at the limit of detection, 3) prolonged shedding, potentially combined with either of the former, or 4) reinfection. The patient with the longest duration between negative rt-PCR and re-positive was enrolled in an observational study to distinguish between these possibilities.

**Figure 1:**
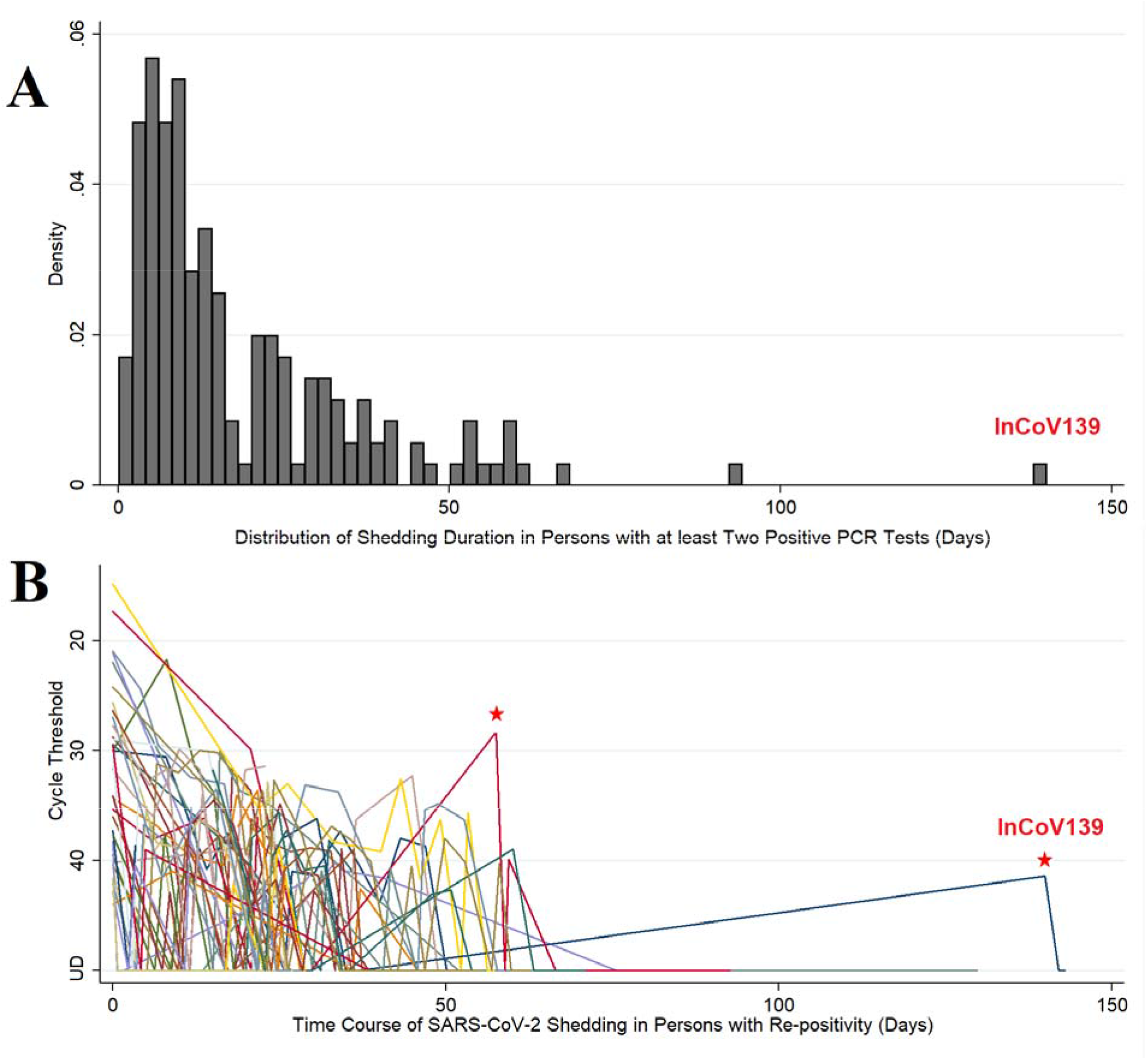
Population viral RNA shedding from patients with COVID-19. Panel A: Distribution of shedding duration in patients who had at least 2 positive SARS-CoV-2 PCR tests. The shedding duration was calculated as the time from first positive sample to last positive sample. In the histogram (n=176), the proportion of patients is plotted as density on the y-axis and shedding duration (in days) is on the x-axis. Panel B: Time course of SARS-CoV-2 shedding in patients (n=43) who had “re-positive” pattern (repeat SARS-CoV-2 PCR positive after negative testing in patients with initially PCR-confirmed COVID-19, i.e. a positive-negative-positive pattern). In the spaghetti plot, semi-quantitative real-time PCR expressed in cycle thresholds (Ct) is plotted on the y-axis and time course in days from first positive to last positive is on the x-axis. Ct is the average result of E & N2 genes except where one target was undetectable and then Ct was set to value of single positive target. Ct range: 14.9 – 44.0. Negative (undetectable) results are set to Ct = 50 for purposes of display. UD = undetectable. Red stars mark possible reinfections due to low CT value at re-positive, or long duration since last positive PCR, respectively.

### Case Study

InCoV139 is a sexagenarian (age between 60 – 69), who resides in a skilled nursing facility (SNF) and has a history of severe emphysema (FEV1 34% predicted) on home oxygen, and hypertension. When hospitalized in early March for severe COVID-19 pneumonia, symptoms included fever, chills, productive cough, dyspnea and chest pain. The patient reported exposure to a SNF employee recently returned from the Philippines with respiratory infection. Auscultation revealed diffuse wheezing and dullness at the left base and chest X-ray showed hyperinflation and bibasilar infiltrates. Unstable atrial fibrillation ensued and was treated with cardioversion and anticoagulation. The patient received treatment with supportive care consisting of supplemental oxygen, steroids and multimodal inhaled therapies for chronic obstructive pulmonary disease. The patient returned to the SNF after testing negative on days 39 and 41 of hospitalization.

InCoV139 remained isolated from family and visitors, interacting only with SNF residents and staff. After moving to a different facility, the patient described exposure to residents at the new facility who were coughing. On day 140 after the first positive SARS-CoV-2 PCR, the patient returned to the ER with dyspnea, reporting 2 weeks of dry cough and weakness. SARS-CoV-2 PCR was repeatedly positive on days 1 and 6 of re-hospitalization (day 14 and day 19 after reinfection date of symptoms onset). Compared to admission in March, the patient was less severely ill in July, by physiologic, laboratory and radiographic parameters, with higher Ct values (**Table 1, Figure S2**). Status returned to baseline after treatment with remdesivir and dexamethasone.

**Table 1:** Clinical Parameters at Peak Illness for COVID-19 Episodes.

| **Parameter:** | **Primary Infection**<br>**(March) \*** | **Reinfection**<br>**(July) \*** |
| --- | --- | --- |
| Vital Signs: |  |  |
| Temperature (°C) | 38.4 | 37.0 |
| Heart Rate (/minute) | 101 | 86 |
| Blood pressure (mmHg) | 156/96 | 143/93 |
| Respiratory Rate (/minute) | 20 | 19 |
| SpO2 (%) on supplemental O<sub>2</sub> rate | 93% on 6 L/min | 94-97% on 3 L/min |
| BMI (kg/m<sup>2</sup>) | 18.7 | 20.4 |
| Laboratory: |  |  |
| Total white blood count (cells/μL) | 16,200 | 6,700 |
| Absolute neutrophil count (cells/μL) | 12,960 | 2,010 |
| Absolute lymphocyte count (cells/μL) | 1,600 | 600 |
| Hematocrit (%) | 39.6% | 42.8% |
| Platelet count (cells/μL) | 290,000 | 240,000 |
| D-dimer (≤0.49 μg/mL) ** | N/A | 0.47 |
| Creatinine (mg/dL) | 1.01 | 1.07 |
| Procalcitonin (≤0.25 ng/mL) ** | 0.15 | 0.08 |
| C-reactive protein (≤5 mg/L) ** | N/A | <3.0 |
| SARS-CoV-2 rt-PCR CT (target 1) † | 22.8 (E) | 43.3 (E) |
| SARS-CoV-2 rt-PCR CT (target 2) † | 26.5 (RdRp) | 39.6 (N2) |

\* Peak day of illness for each COVID-19 episode is given. For primary infection, peak illness occurred in the 1<sup>st</sup> hospitalization (March) on hospital day #5. For reinfection, peak illness occurred in the 2<sup>nd</sup> hospitalization (July) on hospital day #1.
\*\* normal ranges given as indicated
† SARS-CoV-2 qualitative polymerase chain reaction (rt-PCR) cycle threshold (CT) was based on the WHO assay in March (UW Virology) and the Cepheid Infinity in July (LabCorp Seattle).

### Viral Sequencing and Phylogenetic Analysis

Comparison of InCoV139 sequences from March and July revealed 10 high confidence intra-host single nucleotide variants (iSNVs) of which 5 type the March sequence to clade 19B, and 5 type the July sequence to 20A. The InCoV139-March sequence (Genbank: MT252824) shares the canonical mutations (C8782T and T28144C) which define clade 19B and distinguish it from the original clade 19A, Wuhan-Hu-1 reference strain (Genbank: NC_045512.2). InCoV139-March additionally shares C18060T with the first US case WA1 (Genbank: MN985325), which was circulating in Asia and introduced via a traveler returning from Wuhan, China to the Puget Sound area north of Seattle in mid-January.^17^ InCoV139-March diverges from WA1 by 2 other mutations suggesting evolution via community spread in the ensuing 7 weeks from diagnosis of WA1 to diagnosis of InCoV139-March. Conspicuously, the July sequence (InCoV139-July) harbors none of the canonical mutations defining clade 19B and instead shares the canonical mutations defining clade 20A (C3037T, C14408T and A23403G), 1 canonical mutation of clade 20C (G25563T), as well as 1 other 20A mutation. Importantly, present in InCoV139-July (but not in InCoV139-March) is the A23403G mutation, which confers the D614G amino acid change in spike protein, and defines the SARS-CoV-2 strain with greater replicative fitness, introduced separately to the US East Coast via Europe.^18^ As indicated in the phylogeny (**Figure 2**), the iSNVs (**Table S1**) clearly define 2 genetically distinct viruses which evolved separately from a common ancestor in early divergent events.

**Figure 2:**
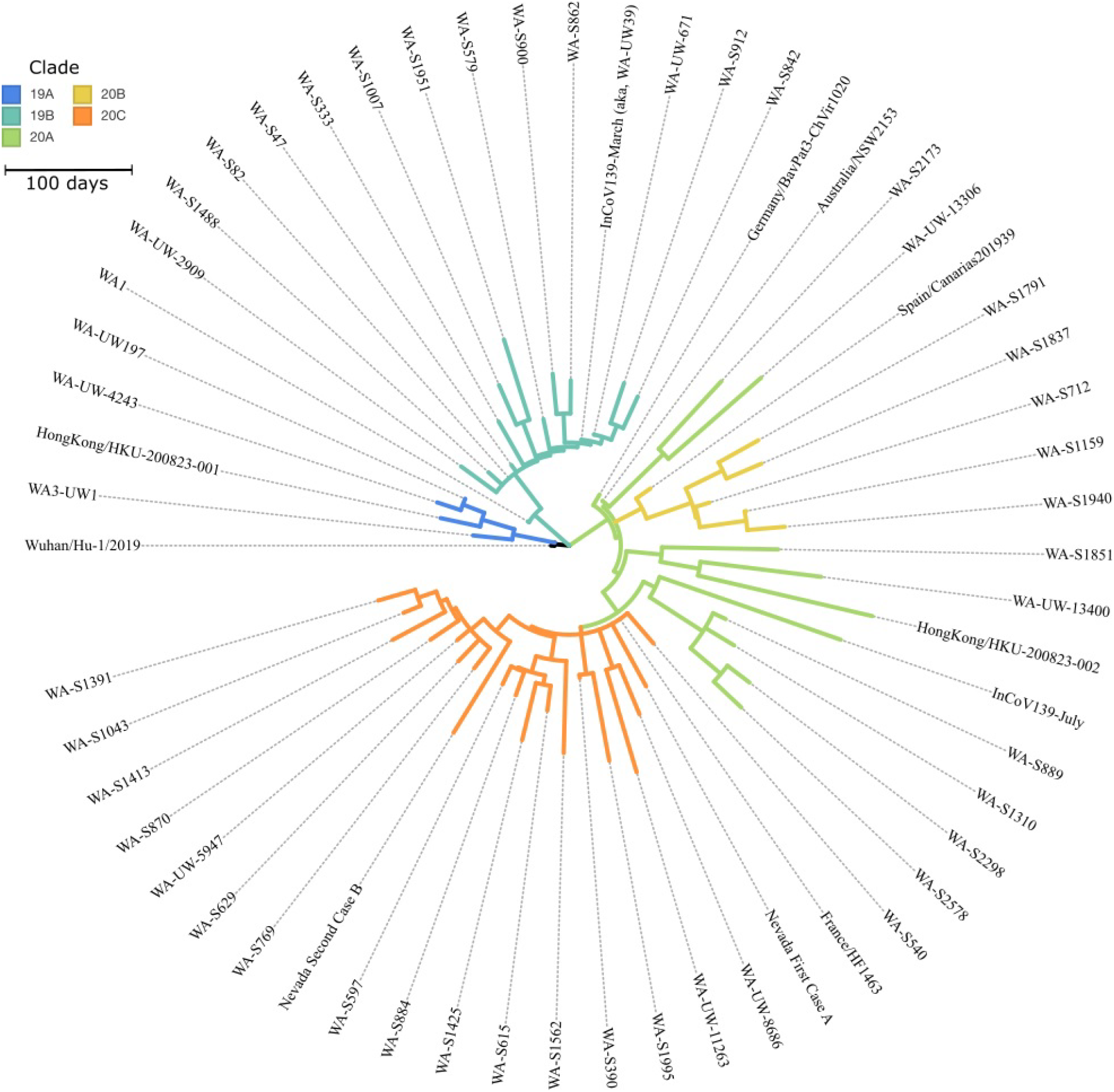
Phylogram of SARS-CoV-2 Isolates in Washington State. Phylogeny of SARS-CoV-2 in Washington State, including contextual sequences, and the pairs of reported reinfection cases from Hong Kong and Nevada. Sequences are colored by clade, as designated by NextClade as follows: 19A: royal blue, 19B: teal, 20A: green, 20B: yellow and 20C: orange. The pair of reinfection cases from InCoV139 (red) are the sample in clade 19B on March 6 from the initial infection and the sample in Clade 20A on July 29 from the reinfection. The initial Hong Kong sample was in Clade 19A and the reinfection sample in 20A; both Nevada samples were in Clade 20C. Sequences are labeled as per the GISAID nomenclature, with edits for readability and information on each sequence is provided (**Table S2**). An interactive version of this tree is included as supplemental material.

### Anti-SARS-CoV-2 Antibody Response

Plasma samples from InCoV139 in July were measured for anti-SARS-CoV-2 antibodies (**Figure 3**). IgG antibodies against RBD, spike and nucleocapsid were detected, with low optical density compared to positive control,^13^ and showed a decreasing trend from day 14 to 42 after reinfection symptoms onset. IgM was weakly positive to spike, but undetectable to RBD and nucleocapsid. IgA specific for spike and nucleocapsid, but not RBD, was detected at low levels on day 14 to 21. Anti-spike and anti-RBD IgA showed a surprising increase by day 42, confirmed in replicate and titration experiments (**Figure S3**). IgG subclass analysis revealed that the patient’s RBD-specific IgG response consisted of low levels of IgG3, without detectable IgG1, despite having both IgG1 and IgG3 specific for spike and nucleocapsid proteins with decreasing trend (**Figure S4**). Antibodies blocking ACE2-RBD binding were undetectable at day 14, suggesting a lack of potentially protective antibodies, and increased by day 42 (**Figure 3**). At day 14 and 42, nAb titers (IC50) were 1:260 and 1:382 against D614 (Wuhan) pseudovirus, and were 1:449 and 1:1168 against a mutated D614G pseudovirus, showing differential increase of nAb to D614G pseudovirus compared to the Wuhan strain (**Figures 3D and S5**).

**Figure 3.**
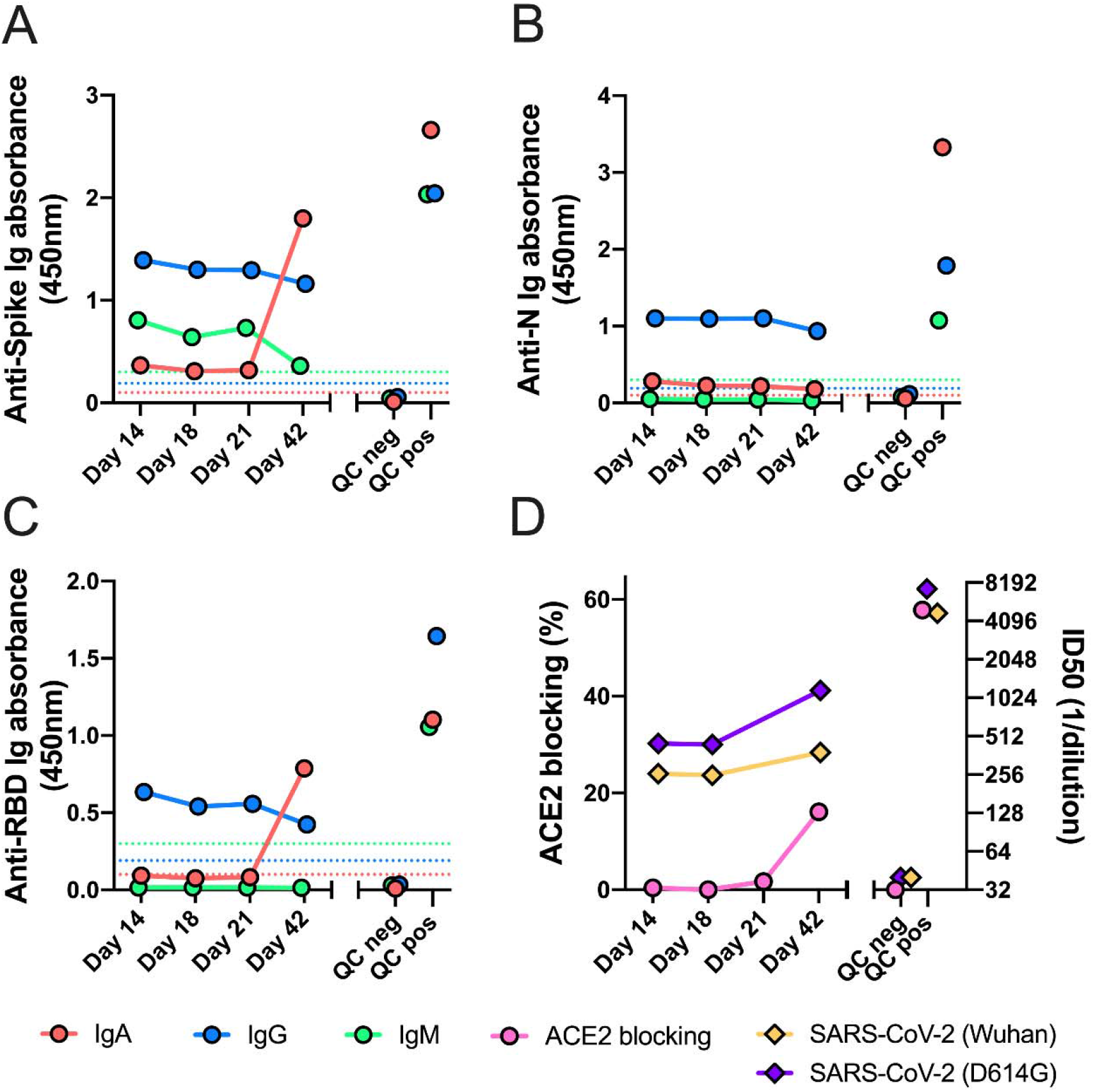
Anti-SARS-CoV-2 serologies and neutralizing antibodies. Plasma samples were analyzed by ELISA at a 1:100 dilution for the presence of IgG, IgA and IgM antibodies binding to the SARS-CoV-2 spike (Panel A), nucleocapsid (Panel B), and RBD (Panel C) antigens. Panel D shows the results of testing for antibodies that block the binding of ACE2 to RBD, carried out with a 1:10 dilution of plasma (left y-axis). Pseudovirus neutralizing antibodies were detected with *in vitro* cell culture assay with D614 (Wuhan) pseudovirus and D614G pseudovirus (right y-axis). For all panels, time on the x-axis indicates days after symptom onset during SARS-CoV-2 reinfection. Plasma pools from SARS-CoV-2 pre-pandemic healthy blood donors and from primary infection COVID-19 patients were used as negative and positive quality control (QC), respectively. The dotted line is the cutoff value for a positive result for each assay, determined as described in the Supplemental Methods. All samples were tested in duplicate wells, and mean OD values are shown. Results are shown from one of two replicate experiments carried out on different days.

### Antibody and B-Cell Receptor Repertoires

B cells were evaluated in peripheral blood at day 14 and 18 after reinfection by NGS of IGH genes of all isotypes (**Figure 4A**). Healthy human peripheral blood shows a predominance of naíve B cells expressing IgM and IgD without somatic hypermutation, and memory B cells with mutated class-switched antibodies. In contrast, the acute response to primary SARS-CoV-2 infection features large polyclonal expansions of recently class-switched, low somatic hypermutation B cells expressing IgG subclasses and, to a lesser degree, IgA subclasses,^19^ as shown in longitudinal samples from an unrelated patient at day 9 (prior to seroconversion) and day 13 (after seroconversion) after primary infection with SARS-CoV-2. In contrast, clones with low somatic hypermutation did not emerge by day 14 or 18 after reinfection in patient InCoV139 (**Figure 4A**). Parallel analysis by single-B cell immunoglobulin sequencing revealed elevated frequencies of IgA-expressing B cells, particularly IgA2-expressing cells (**Figure 4B**).

**Figure 4.**
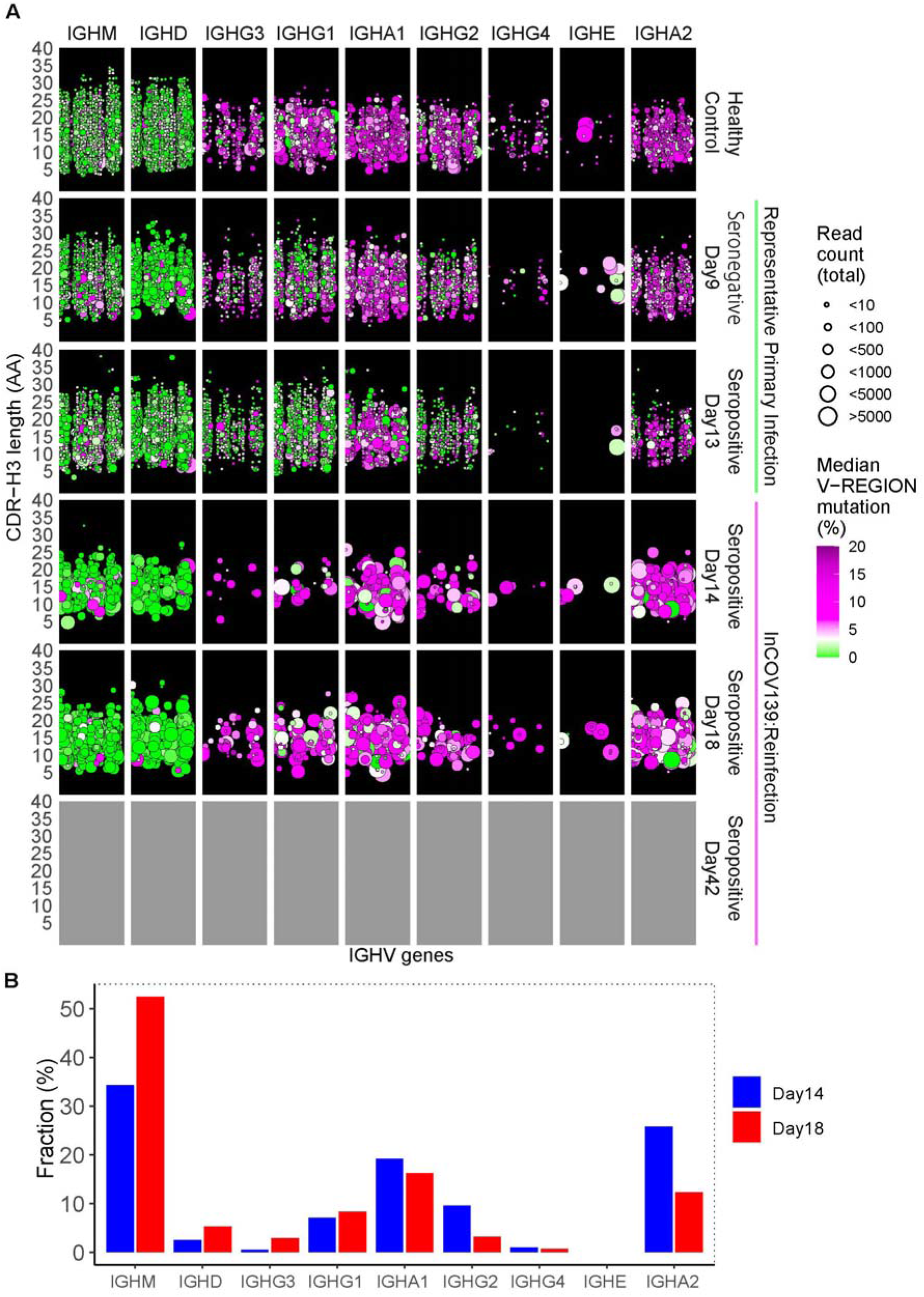
B cell repertoire responses. Panel A: Peripheral blood B cell IGH gene repertoires from peripheral blood mononuclear cell RNA. Three individuals were sampled: a representative healthy control (top row); a patient with representative primary SARS-CoV-2 infection at day 9 and 13 post-onset of symptoms (highlighted by green bar); and reinfected patient at day 14, 18 and 42 (pending) post-onset of symptoms (highlighted by pink bar). Serostatus and days post symptoms onset are given on the right y-axis. Columns indicate the class of each IGH sequence with the IGHV gene indicated on the x-axis. The left y-axis indicates CDR-H3 length in amino acids (AA). Dots indicate B cell clonal lineages. Point color indicates median IGHV somatic hypermutation frequency for each clone, and point size indicates the number of reads in the clone. Points are jittered to decrease over-plotting. Panel B: The bar plot summarizes single-B cell transcriptome data indicating the antibody isotype expressed by B cells in the reinfected patient’s blood, plotted as the frequency of usage of each isotype.

## Discussion

We present a case of SARS-CoV-2 reinfection and perform extensive characterizations of antibody and B cell responses. Our data suggest an initial benchmark to begin understanding the correlates of humoral immunity required to prevent reinfection. Understanding such correlates can aid in planning the re-opening of society as some persons are likely to be at risk for reinfection due to waning antibody-mediated immunity. While vaccine development programs are in full swing, protective levels of total anti-spike antibodies or nAbs are still unknown.

Molecular evidence for reinfection in our patient is strong. At initial infection during the early outbreak in Seattle, sequences of community circulating viruses had low diversity, and were derived from a founder virus introduced to the US some 7 weeks earlier.^17^ The spike variant D614G from Europe has now taken over as the predominant circulating strain.^18^ The time course of InCoV139’s two infections overlaps with the transition in Seattle to the newer D614G strain,^20^ supporting reinfection as opposed to intra-host evolution.

The case patient had anti-SARS-CoV-2 IgG antibodies in the first weeks after reinfection, but notably, levels of anti-RBD IgG were relatively low, with no evidence of blocking antibodies to the RBD-ACE2 complex. ACE2 blocking increased only slightly by day 42, likely due to IgA antibodies. In the B cell repertoire, new clones do not emerge by day 18 after reinfection, lending support to immune recognition of prior infection or suggesting a deficiency in the developing response to the reinfection. While we do not know the nAb titers immediately at the time of reinfection, by day 14 after reinfection, nAb levels were comparable to those observed after boosted vaccination.^21^ By day 42 nAb response showed a 1.5-fold increase to Wuhan pseudovirus, and a 2.6-fold increase to D614G pseudovirus. Taken together, these findings suggest that poorly developed or waned antibodies against the D614 virus formed after primary infection in March were not protective against reinfection with the D614G spike variant acquired in July. These results could have important implications for the success of vaccine programs based on the Wuhan strain.

Fortunately for our patient, the reinfection was more mild than was the primary infection, in contrast to the Nevada case.^11^ This case report provides an initial in-depth assessment of humoral immune responses during reinfection. Furthermore, the humoral immunity levels provide a starting point which describe a benchmark which is shown not to be protective against reinfection. Larger case series of reinfection patients or follow-up experience after vaccination studies will be needed to more thoroughly evaluate correlates of immune protection against SARS-CoV-2.

## Methods

### Patient Population & PCR Testing

“Re-positivity” was defined as repeat SARS-CoV-2 PCR positive after negative testing in patients with initially PCR-confirmed COVID-19. To understand the duration of shedding and phenotypes of re-positivity, we analyzed a database of all SARS-CoV-2 PCR testing for patients with nasopharyngeal samples sent from the emergency departments or hospitals of Swedish Health System in Seattle, WA. Semi-quantitative real time polymerase chain reaction (rt-PCR) testing reported as cycle thresholds (Ct) were performed on the Xpert® Xpress SARS-CoV-2 test on the GeneXpert Infinity (Cepheid, Sunnyvale, CA). Hospital policies discouraged unnecessary testing, but the decision to test was left to individual providers. Retesting was often requested by congregate living facilities prior to receiving patients following hospitalization. Discontinuation of transmission-based isolation synchronized with the CDC interim guidance.^22^ Descriptive statistics were performed on population shedding dynamics.

### Virologic and Immunologic Analyses

Viral sequencing in March was performed via rapid metagenomic next-generation sequencing (NGS),^23^ and in July was modified from the multiplexed PCR amplicon NGS method using the ARTIC V3 primers.^24^ SARS-CoV-2 clade designations and phylogenetic analyses were produced using NextStrain.^25^ Serological testing used enzyme-linked immunosorbent assay (ELISA) for anti-spike, anti-RBD, and anti-N IgG, IgM and IgA antibodies, as well as a functional assay for antibodies that block binding of RBD to an ACE2 fusion protein.^13^ Functional nAbs were measured with a cell-culture based assay using pseudoviruses containing either the D614 or the G614 epitopes in spike.^21^ Immunoglobulin heavy chain (IGH) genes expressed by peripheral blood B cells were sequenced with amplicon libraries produced for each isotype,^19^ and paired IGH and light chain sequences were determined with single B cell transcriptome analysis.^19^ All assays are described in depth (Supplemental Methods).

### Ethical Approval

The Providence St. Joseph’s Health IRB approved the study (STUDY2020000175). Informed consent was waived for use of residual samples and population clinical data (STUDY2020000143).

## Data Availability

All data will be made public upon publication of the results in a peer-reviewed journal, as per our funding agreement with BARDA.

## Funding Statement

This project has been funded with support from the Murdock Charitable Trust (JRH), the Jeff and Liesl Wilke Foundation (JRH), the Swedish Medical Center Foundation (JDG), the Coulter Foundation (SDB), and federal funds from the National Institutes of Health, National Institute of Allergy and Infectious Diseases, 3R01AI141953-02S1 (JRH), and from the Department of Health and Human Services; Office of the Assistant Secretary for Preparedness and Response; Biomedical Advanced Research and Development Authority, under Contract No. HHSO100201600031C, administered by Merck & Co (JRH, JDG).

## Competing Interest Statement

JDG and JRH received U.S. federal funding from Department of Health and Human Services; Office of the Assistant Secretary for Preparedness and Response; Biomedical Advanced Research and Development Authority, under Contract No. HHS0100201600031C, which is administered by Merck & Co. JDG performed contracted research with Gilead Sciences, Inc. ATS is a scientific founder of Immunai and receives research funding from Arsenal Biosciences. All other authors report no competing interests. The sponsor had no role in any stage of the study, including manuscript preparation, but reviewed the manuscript prior to submission.

## Author Contributions

Study Concept: JDG, SDB and JRH; Resources/funding: JDG, SDB and JRH; Clinical care: JDG, JJ, GPP; Methods: JDG, KW, JCR, KC, CJP, ALG, SDB and JRH; Performed experiments or collected data: JDG, KW, KR, SCAN, JCR, SNN, FY, OFW, KEY, JYL, TW, CJP, IL, SF, YS, JH, KMM, PMM, JAW, HAA, ALG, ATS; Figures / data analysis: JDG, KW, JCR, KC, CJP, KR, SCAN, SDB; Interpreted analyses: JDG, KW, JCR, SNN, ALG, KC, CJP, KR, SCAN, SDB, JRH; Administrative support: JAW, HAA, KMM; Literature search: JDG, JJ, WRB, STN, JSP; Wrote the first draft: JDG; Revised the manuscript and approved the final version: all authors.

## Data Availability Statement

The full data is available upon request. Code used to generate analyses is available upon request. Viral sequences will be deposited into GenBank prior to publication.

## Notes

### Author Declarations

The Providence St. Joseph Health IRB approved the study (STUDY2020000175). Informed consent was waived for use of residual samples and population clinical data (STUDY2020000143).

